# Refractive error prevalence in urban Vietnamese children: a 2021 to 2024 school health analysis

**DOI:** 10.64898/2026.09.08.26362493

**Authors:** Khoi Pham Minh Tran, Nhan Thi Ho

**Affiliations:** College of Health Sciences, VinUniversity, Hanoi, Vietnam; Pediatric Department, Vinmec Times City, Vinmec International Hospital, Hanoi, Vietnam; Research Management Department, Vinmec International Hospital, Hanoi, Vietnam

**Keywords:** refractive error, prevalence, Vietnamese, children, urban, school health

## Abstract

**Background:** Refractive error is a leading cause of childhood visual impairment worldwide, yet longitudinal, multi-city data from Vietnam remain scarce, especially for children younger than school age. We aimed to describe the prevalence and secular trends of refractive error in a large urban Vietnamese school health program.

**Methods:** We conducted a retrospective analysis of repeated cross-sectional school health screening data collected from 2021 to 2024 across three major Vietnamese cities (Hanoi, Ho Chi Minh City, and Hai Phong), among children aged 3 to 18 years. The dataset included 141,339 examinations, with a Hanoi sub-cohort of 19,788 examinations that had quantitative autorefractor measurements. We estimated crude prevalence and adjusted prevalence ratios using Poisson regression.

**Results:** The prevalence of any refractive error stayed high throughout the study period, ranging from 53.9% to 57.1% (P for trend < 0.001), and was higher in Hai Phong (68.4%) and Hanoi (61.6%) than Ho Chi Minh City (35.9%), higher in girls than boys (58.5% vs 53.1%), and higher with older age (P < 0.001). In the Hanoi sub-cohort, confirmed myopia prevalence nearly tripled, from 3.7% in 2022 to 11.0% in 2024 (P < 0.001), while hyperopia declined.

**Conclusions:** Refractive error is highly prevalent among urban Vietnamese schoolchildren, and confirmed myopia appears to be rising rapidly at younger ages. These findings support closer surveillance and earlier school-based intervention in Vietnam and similar settings.

## INTRODUCTION

Refractive error, mainly myopia, hyperopia, and astigmatism, is the leading cause of visual impairment in children and the second leading cause of vision loss overall, accounting for roughly 43% of all visual impairment worldwide^1^. A global meta-analysis of studies from 1990 to 2016 estimated the pooled prevalence of myopia, hyperopia, and astigmatism in children at 11.7%, 4.6%, and 14.9%, with astigmatism the most common overall and prevalence varying considerably by region^1^. A review of 80 studies of school-age children worldwide reported cycloplegic myopia prevalence reaching 60% in Asian populations compared with 40% in Europe, pointing to low outdoor time and intensive near work as the main modifiable risk factors^2^. Global Burden of Disease 2021 estimates put refraction disorders at nearly 160 million prevalent cases that year. The case numbers keep rising, and the burden stays concentrated in low– and middle-income regions^3^.

East and Southeast Asia carry a disproportionate share of this burden. Meta-analytic data show myopia prevalence among children rose from 10.4% in 1993 to 34.2% in 2016 globally, with East Asian populations showing the steepest trajectory, roughly a 23% increase per decade, reaching 69% by age 15 compared with under 6% in African children of the same age^4^. Within Southeast Asia, prevalence varies enormously from country to country. Schoolchildren in Lao PDR show a myopia prevalence below 1%, with the great majority retaining normal unaided vision^5^, while in neighboring Cambodia, prevalence differs sharply between urban Phnom Penh (13.7%) and rural Kandal province (2.5%)^6^. Whether this patchwork points toward an approaching epidemic similar to East Asia’s is still debated. Most current evidence suggests prevalence at the end of schooling in Southeast Asia remains modest, although emerging young-adult data from Thailand and Indonesia hint it may be approaching epidemic levels, and regional education indicators do not yet clearly signal one^7^.

Vietnam sits somewhere in the middle of this regional gradient. A recent meta-analysis of Vietnamese studies estimated an overall pooled refractive-error prevalence of 37.6% among schoolchildren, with myopia accounting for about 29%, equivalent to an estimated 6 to 8 million affected children nationally, and prevalence significantly higher among girls and in urban settings^8^. The same evidence points to a rapid secular rise, with secondary-school myopia prevalence climbing from 13.1% in 2009 to 33.7% by 2014^8^. Smaller studies tell a similar story. In Ba Ria Vung Tau province, uncorrected refractive error accounted for most vision impairment among secondary schoolchildren, with myopia present in about one in five^9^, while in rural Nghe An province, myopia rose from 10.5% in grade six to 17.7% in grade nine and was linked to maternal education, parents wearing spectacles, close reading distance, and less outdoor time^10^.

Despite this growing evidence, some important gaps remain. Existing Vietnamese data come almost entirely from single-province, single-time-point surveys of school-age children, so little is known about multi-year trends within a defined population, or about how refractive error develops in preschool-aged children, and a recent meta-analysis noted wide confidence intervals across pooled estimates and called for standardized, longitudinal protocols^8^. Much of the regional literature also focuses heavily on myopia, while hyperopia and astigmatism get comparatively little attention, even though astigmatism is actually the most common refractive error in children globally^1^. Evidence from China shows the age of myopia onset has been shifting earlier over time, from a mean of 10.6 years in 2005 to 7.6 years in 2021^11^, a shift that matters clinically since children with onset at 7 or 8 years have a 53.9% probability of developing high myopia in adulthood, compared with only 1.3% for onset at 12 years or later^12^. Vietnam, and Southeast Asia more broadly, still lacks the longitudinal, multi-year cohort data needed to tell whether a similar shift is happening here too.

Our study performed retrospective analysis of a repeated cross-sectional annual school health screening program on children aged 3 to 18 years from a private school system across three major Vietnamese cities, Hanoi, Ho Chi Minh City, and Hai Phong, from 2021 to 2024, combining standardized visual-acuity and refractive-error assessment with a quantitative autorefraction sub-cohort in Hanoi. With more than 140,000 examinations across four years, our study aims to describe the prevalence of refractive error, overall and by type (myopia, hyperopia, astigmatism, and anisometropia), among children screened in three major Vietnamese cities between 2021 and 2024, to characterize secular trends across the study period, and to identify demographic and geographic correlates of refractive error, including age, sex, city, and school level. By drawing on a large, multi-city, multi-year urban cohort spanning early childhood through adolescence, this study aims to provide contemporary evidence to help guide school-based vision-screening policy and refractive error prevention and control strategies in Vietnam and similar countries.

## METHODS

### Study Design, Setting and Participants

Our study analyzed retrospective de-identified annual school health screening data of a private school system collected consecutively from 2021 to 2024 across three major metropolitan areas: Hanoi, Ho Chi Minh City, and Hai Phong. The study was approved by Vinmec Ethical Committee (approval number 0231/2024/CN/HDDD VMEC). Informed consent and written informed consent from the parent/guardian of participant under 18 years of age was waived for this study because this study performed retrospective analysis of de-identified data. The identified data used for this study was accessed on November 1, 2025 and the authors had no access to information that could identify individual participants during or after data retrieval.

### Participants

The study population included children and adolescents ranging from 3 to 18 years of age. To facilitate age-based comparisons, participants were categorized into four distinct school levels: Kindergarten for ages 3 to 5, Primary for ages 6 to 11, Lower secondary for ages 12 to 14, and Upper secondary for ages 15 to 18. During the data cleaning phase, we excluded records belonging to individuals outside this target age range. We additionally excluded examinations missing essential demographic information including sex, examination year, or study site. Finally, records with missing or indeterminate final refractive error classifications were removed from the primary analysis. Following these exclusions, the primary analytic dataset comprised 141,339 valid eye examinations. Furthermore, we established a dedicated sub-cohort of 19,788 examinations from the Hanoi site, which contained quantitative autorefractor measurements, allowing for highly detailed dioptric evaluations.

### Variables and Definitions

Our primary clinical outcome was the presence of any refractive error, which was determined by the diagnostic classification recorded during the school screening. We also assessed impaired visual acuity, strictly defining it as a Snellen visual acuity of less than 6/12 in the worse eye.

Within the Hanoi autorefractor sub-cohort, we categorized specific refractive error types relying on precise quantitative data. Myopia was defined as a spherical equivalent refractive error of – 0.50 diopters or less in either eye^13^. We further classified moderate to high myopia using a threshold of a spherical equivalent of –3.00 diopters or worse. Hyperopia and anisometropia were identified based on specific autorefractor screening codes. Astigmatism was formally defined as a cylinder error measuring 0.75 diopters or greater.

### Statistical Analysis

We summarized all baseline participant characteristics using standard descriptive statistics. Categorical variables such as sex, age group, school level, and city were reported using frequencies and percentages. Continuous variables, notably age and spherical equivalent measurements, were presented as means accompanied by their standard deviations. We calculated the crude prevalence for all refractive error outcomes and reported them alongside their respective 95% Wilson confidence intervals.

We estimated adjusted prevalence ratios adjusting for potential demographic confounders and account for clustering effects at the city level using Poisson regression models specifying robust standard errors known as HC3^14^. These multivariable regression models were comprehensively adjusted for the continuous examination year, city, sex, and chronological age. We performed all data wrangling and statistical analyses using R software version 4.5.1^15^. A two-sided p-value of less than 0.05 was adopted as the threshold for statistical significance across all tests.

## RESULTS

### Study population

A total of 159,181 school health examination records collected between 2021 and 2024 were initially identified. After excluding 6,309 examinations from children aged <3 or >18 years and 16 additional records that did not meet eligibility criteria, 152,856 examinations were eligible for analysis. A further 11,517 examinations with missing or indeterminate refractive error (RE) assessments were excluded, leaving 141,339 examinations in the primary analytic dataset. Quantitative refractive measurements obtained using autorefractors were available for a Hanoi sub-cohort comprising 19,788 examinations (**Figure 1**).

**Figures 1.**
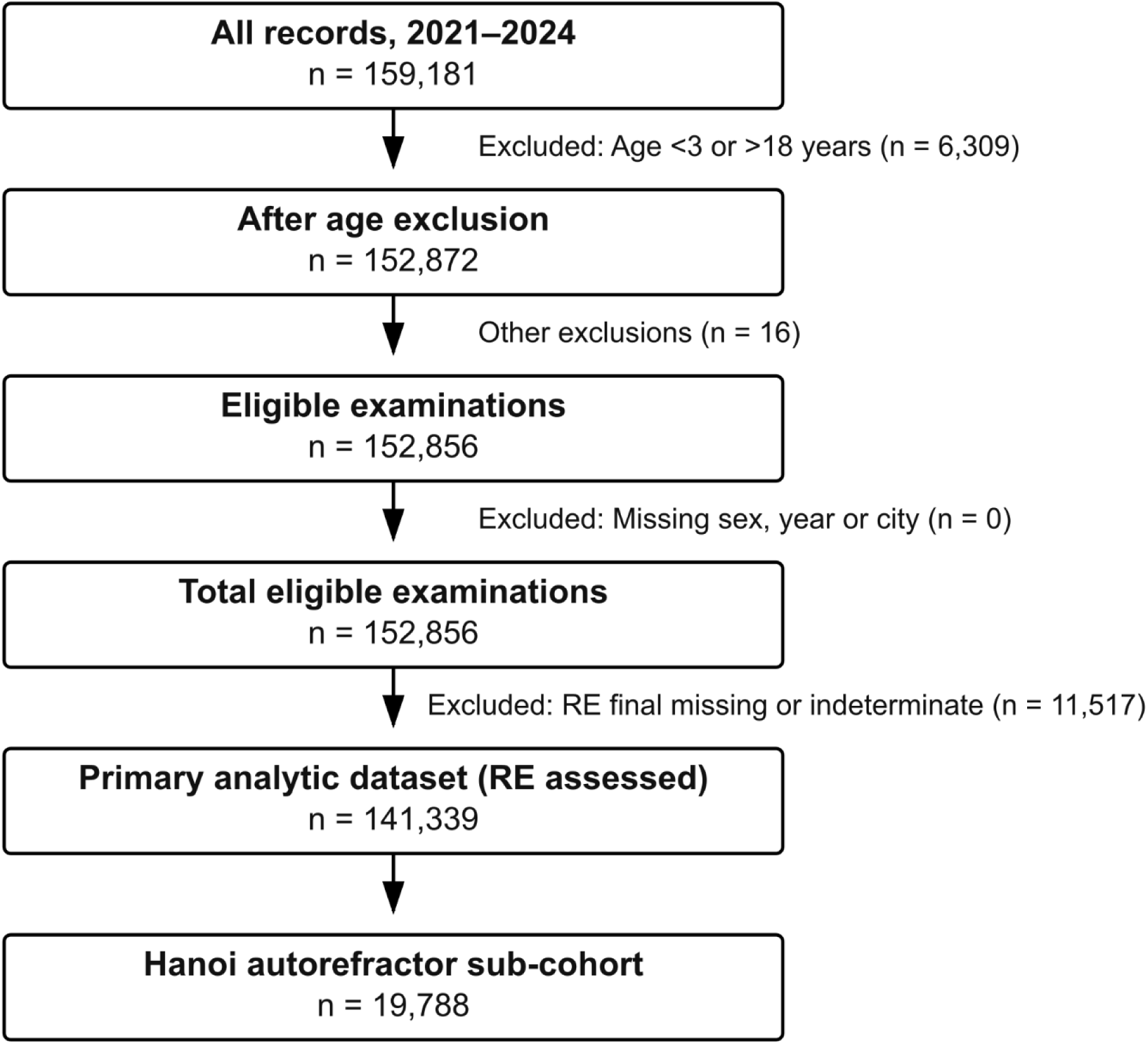
Flow of study participants and examinations included in the analysis, 2021 to 2024. Flow diagram showing the selection of study examinations from the original school health database. Examinations from children aged <3 or >18 years were excluded, followed by exclusions for other reasons and examinations with missing or indeterminate final refractive error (RE) classification. The final analytic dataset comprised 141,339 examinations, including a Hanoi autorefractor sub-cohort of 19,788 examinations used for quantitative refractive error analyses. RE = refractive error.

### Participant characteristics

The characteristics of the eligible study population are summarized in **Table 1**. Among the 152,856 eligible examinations, 52% were from boys and 48% from girls. The mean age was 9.1 ± 3.9 years (range = 3 to 18 years). Approximately half of all examinations were performed in children aged 6 to 11 years (50.0%), followed by those aged 3 to 5 years (22%), 12 to 14 years (17%), and 15 to 18 years (11%). Primary school students accounted for the largest proportion of examinations (42%), while 71% of examinations were conducted in Hanoi, 22% in Ho Chi Minh City, and 6.9% in Hai Phong. Although the distribution of demographic characteristics differed statistically across calendar years (all *P* < 0.001), these differences likely reflected changes in the composition of the school health program rather than large demographic shifts (**Table 1**).

**Table 1.**
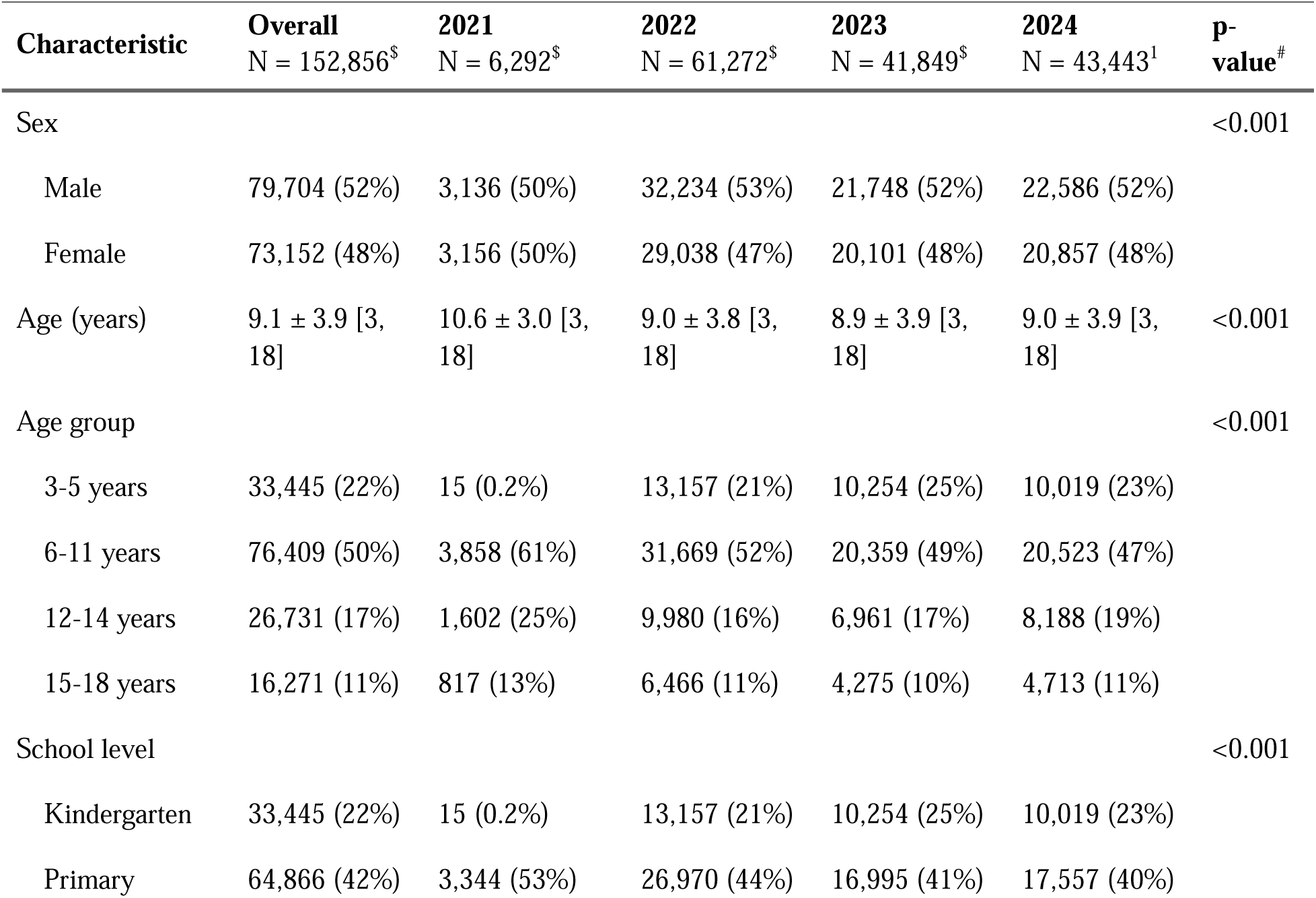
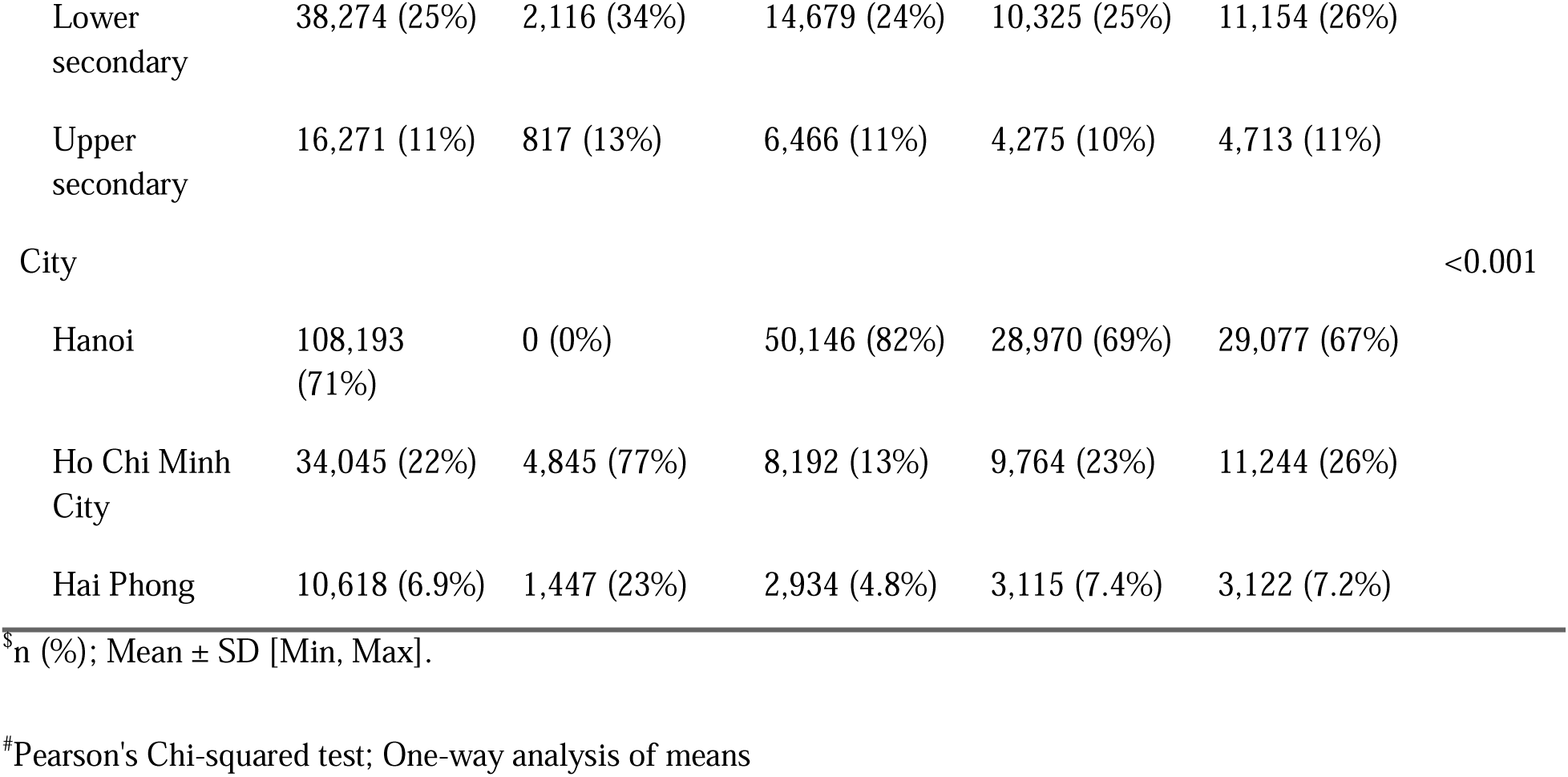
Characteristics of participants by year of examination, 2021 to 2024.

### Refractive error (RE) prevalence

Among examinations with complete RE assessment, the crude prevalence of any refractive error remained consistently high throughout the study period, with overall prevalence of 56.2% in 2021, 57.1% in 2022, 53.9% in 2023, and 55.5% in 2024 (**Table 2**). Despite these modest annual fluctuations, the Cochran-Armitage test indicated a significant temporal trend (*P* < 0.001). In contrast, the prevalence of impaired visual acuity varied between 52.7% and 62.1%, with no evidence of a monotonic temporal trend (*P* = 0.832) (**Table 2**).

**Table 2.**
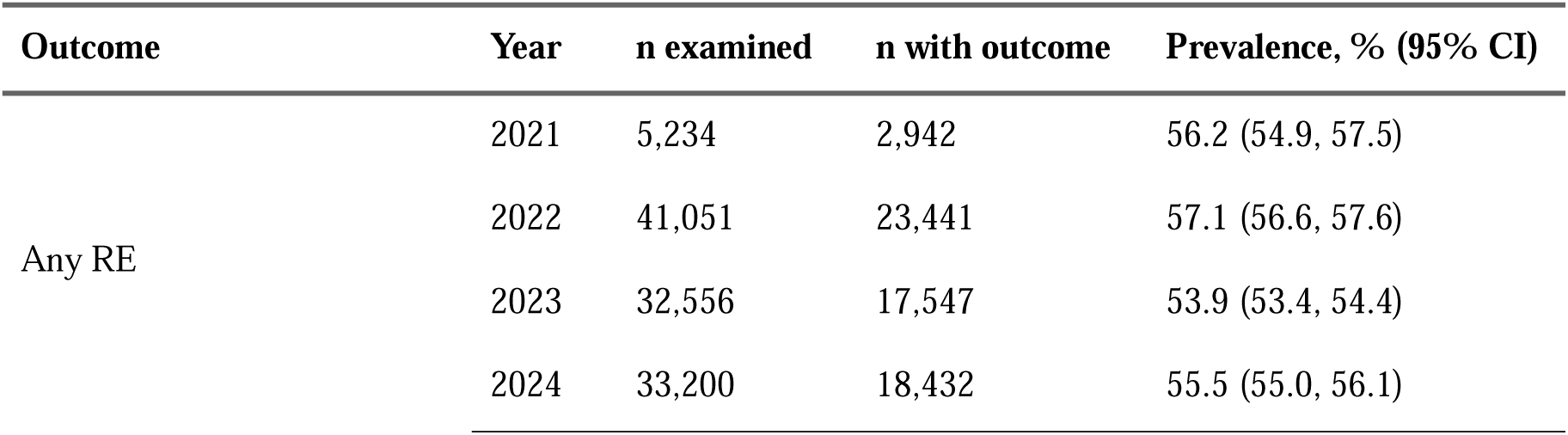
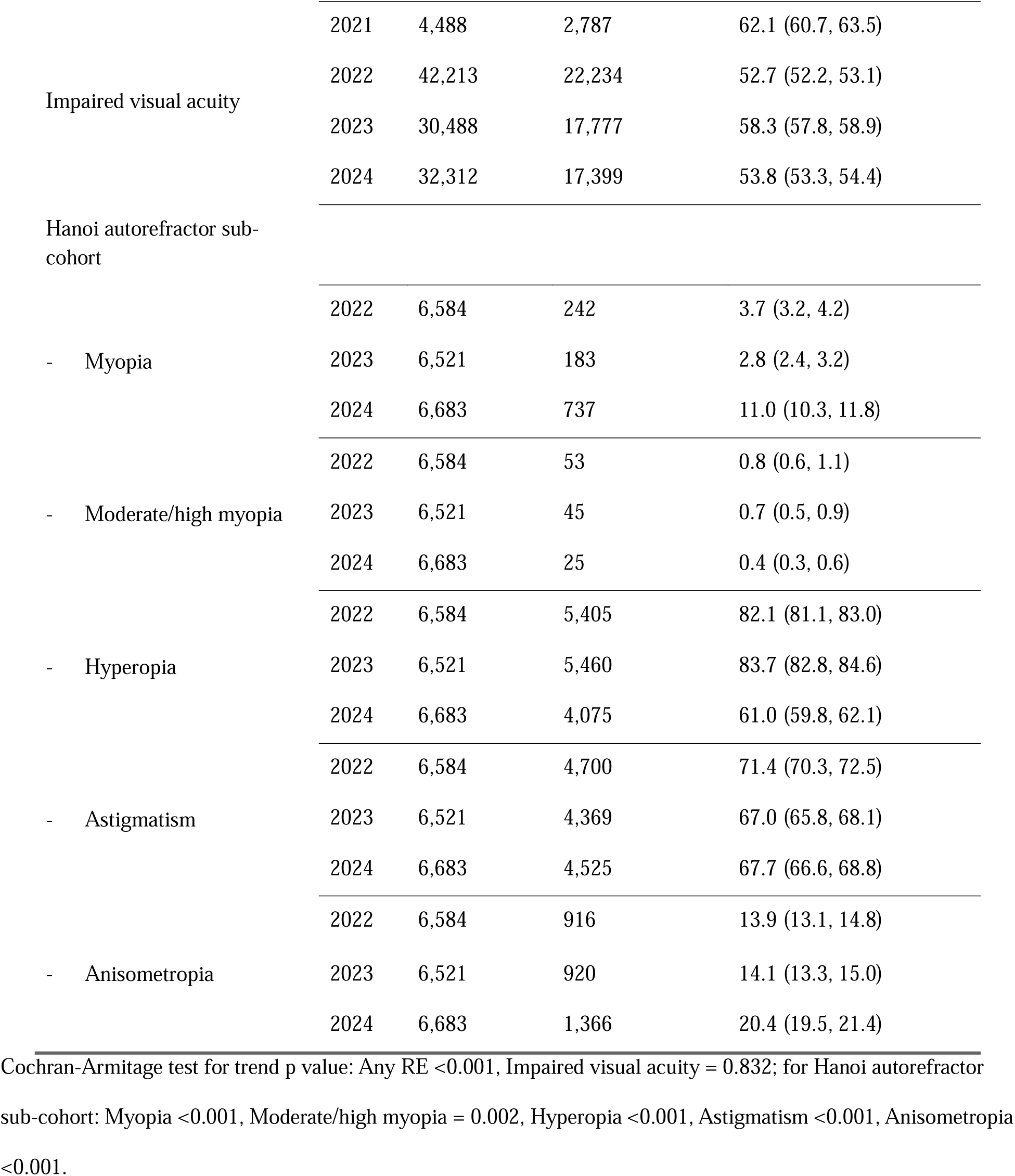
Crude prevalence of refractive errors by year of examination, 2021 to 2024.

Substantial geographical variation in RE prevalence was observed (**Figure 2**, **Table 3, Table S2**). Across all study years, Haiphong exhibited the highest prevalence of refractive error (68.4%), whereas Ho Chi Minh City showed considerably lower prevalence (35.9%) compared to 61.6% in Hanoi (**Table S2**). After adjustment, compared with Hanoi, children from Ho Chi Minh City had substantially lower RE prevalence (aPR= 0.58, 95% CI= (0.57, 0.59)), whereas children from Hai Phong had higher prevalence (aPR= 1.08, 95% CI= (1.06, 1.09)) (both *P* < 0.001) (**Table 3**).

**Figure 2.**
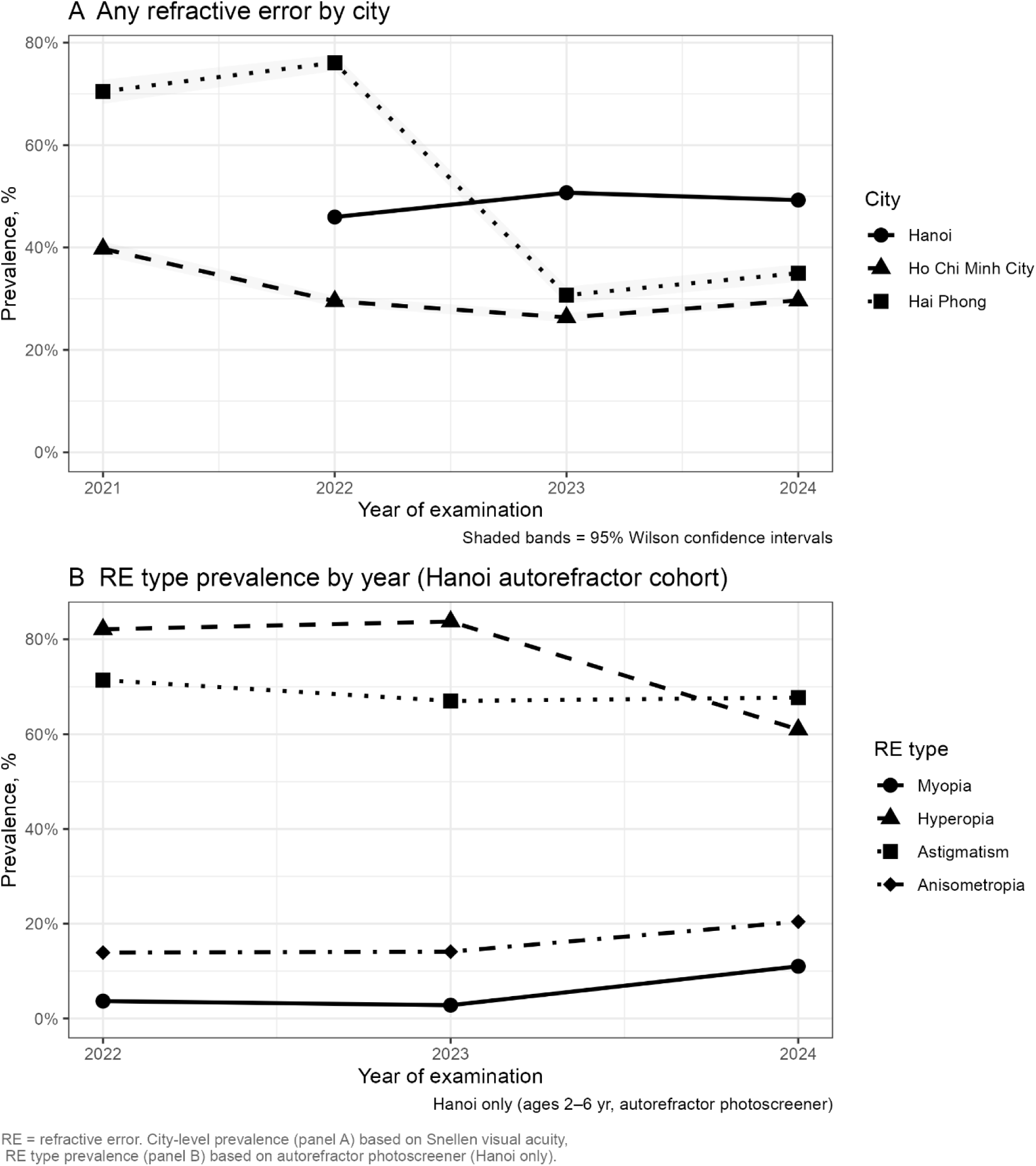
Refractive error prevalence by cities and calendar year. (**A**) Annual prevalence of any refractive error according to city based on school vision screening. Points indicate annual prevalence estimates and shaded bands represent 95% Wilson confidence intervals. **(B)** Annual prevalence of specific refractive error types in the Hanoi autorefractor sub-cohort. Estimates were derived from autorefractor measurements. RE = refractive error.

**Table 3.**
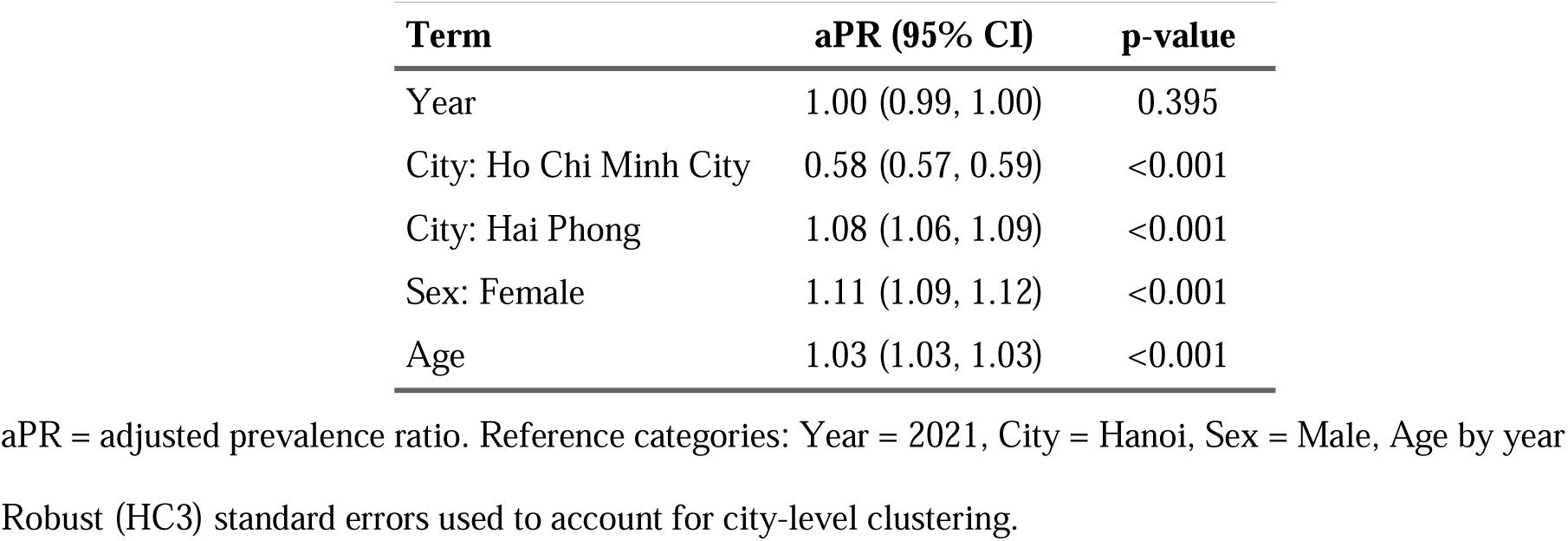
Adjusted prevalence ratios (aPR) for any refractive error from Poisson regression with robust standard errors.

| Term | aPR (95% CI) | p-value |
| --- | --- | --- |
| Year | 1.00 (0.99, 1.00) | 0.395 |
| City: Ho Chi Minh City | 0.58 (0.57, 0.59) | <0.001 |
| City: Hai Phong | 1.08 (1.06, 1.09) | <0.001 |
| Sex: Female | 1.11 (1.09, 1.12) | <0.001 |
| Age | 1.03 (1.03, 1.03) | <0.001 |

aPR = adjusted prevalence ratio. Reference categories: Year = 2021, City = Hanoi, Sex = Male, Age by year
Robust (HC3) standard errors used to account for city-level clustering.

The heat map further demonstrated a clear age-related increase in RE prevalence, with the highest prevalence occurring among older school-aged children, particularly during adolescence (**Figure 3).** Adjusted Poison regression showed that age is independent factor associated with increased refractive error prevalence (aPR = 1.03, 95% CI =1.03, 1.03) **(Table 3**, **Figure 4**). **Figure 5** illustrates the distribution of examination numbers and refractive error prevalence according to age and calendar year. The largest numbers of examinations were performed among primary school-aged children. Refractive error prevalence increased progressively with age, particularly from late primary school onwards, from 32.9% in primary school to 77.8% in upper secondary school (**Table S2**).

**Figure 3.**
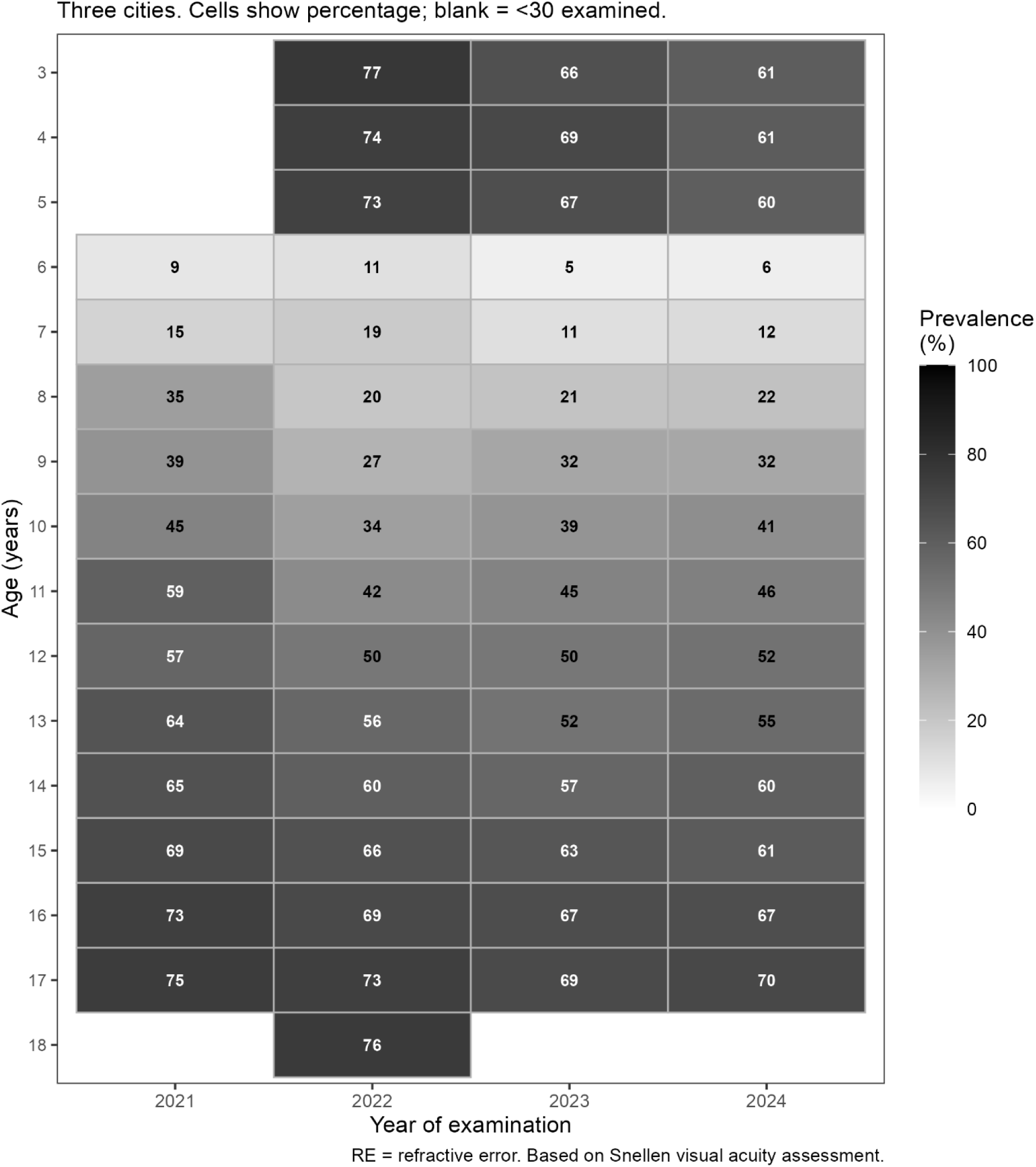
Heat map of refractive error prevalence according to age and calendar year, 2021 to 2024. Each tile represents the prevalence (%) of any refractive error among children of a given age during a calendar year. Color intensity indicates prevalence, with darker colors representing higher prevalence. Cell labels show observed prevalence percentages. Cells with fewer than 30 examinations were suppressed. Estimates were based on school vision screening. RE = refractive error.

**Figure 4.**
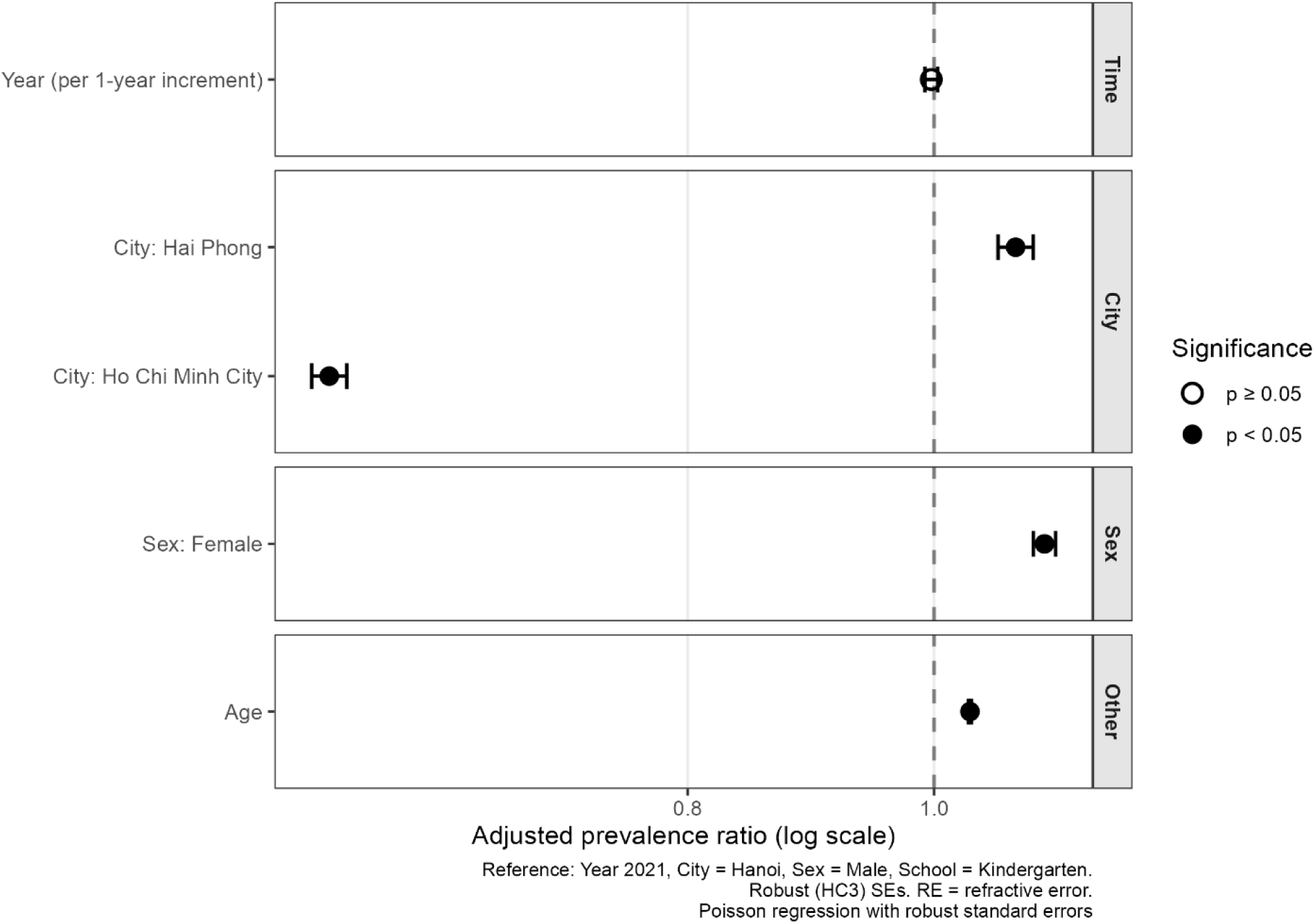
Adjusted prevalence ratios for factors associated with refractive error. Forest plot showing adjusted prevalence ratios (aPRs) and 95% confidence intervals from multivariable Poisson regression with robust standard errors. The vertical dashed line indicates the null value (aPR = 1. Filled symbols indicate statistical significance (P < 0.05), whereas open symbols indicate non-significant associations. Reference categories are shown in the Methods section (or specify them in the figure if preferred). RE = refractive error; aPR = adjusted prevalence ratio.

**Figure 5.**
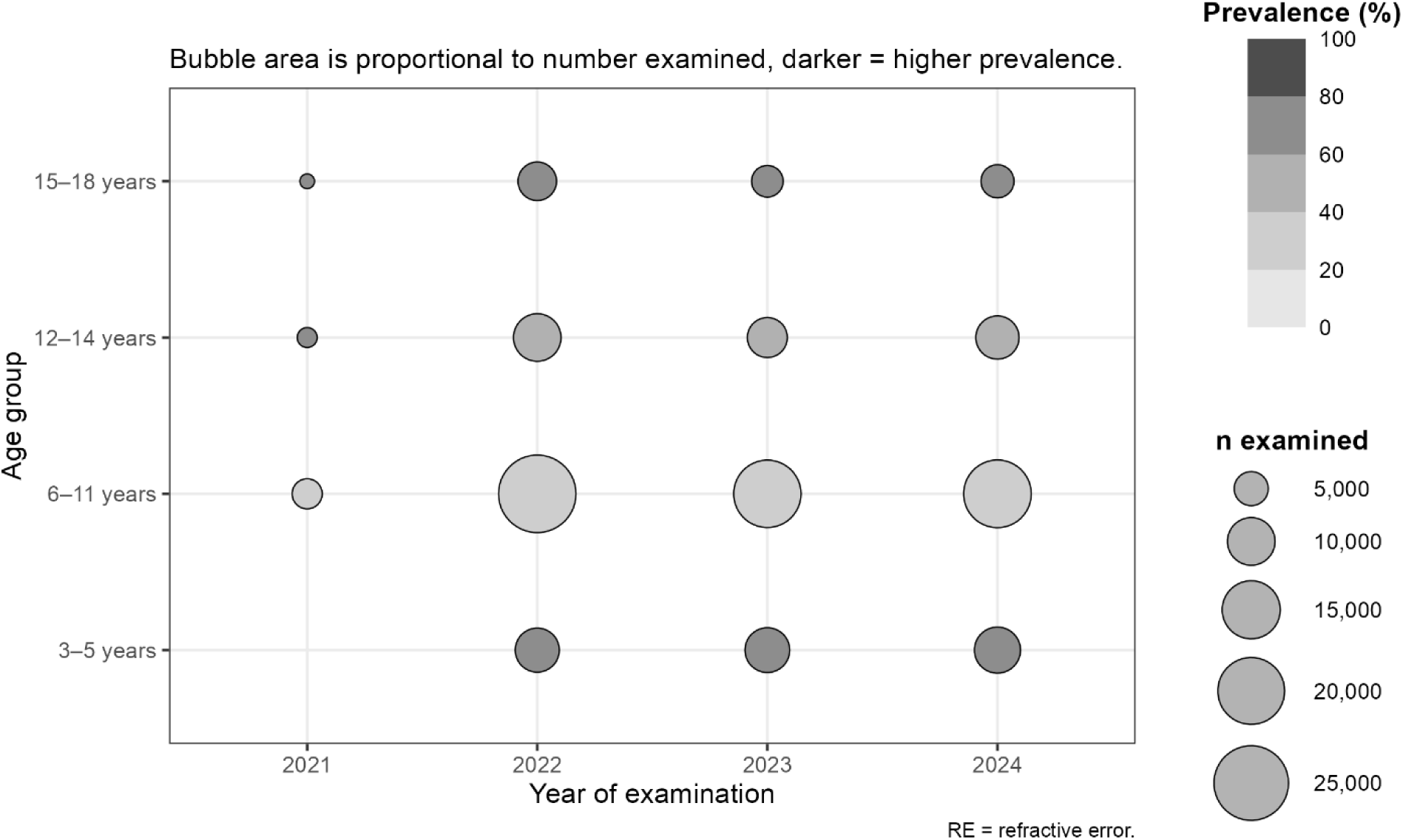
Distribution of sample size and refractive error prevalence according to age group and calendar year. Bubble plot showing the number of examinations and prevalence of any refractive error by age group and year. Bubble area is proportional to the number of examinations, while color intensity represents refractive error prevalence. RE = refractive error.

Sex-stratified analyses showed consistently higher prevalence of any refractive error among girls than boys (58.5% vs. 53.1%) throughout the study period, whereas impaired visual acuity was more common among boys (57.0% vs. 52.8%) (**Figure 6**, **Table S2**). Although prevalence fluctuated modestly between calendar years in both sexes, the temporal patterns were broadly parallel, indicating that the higher prevalence observed among girls persisted across all survey years rather than being driven by a single calendar year. These findings are consistent with the multivariable analysis demonstrating female sex as an independent factor associated with increased refractive error prevalence (aPR =1.10, 95% CI =1.09, 1.12) (**Table 3**).

**Figure 6.**
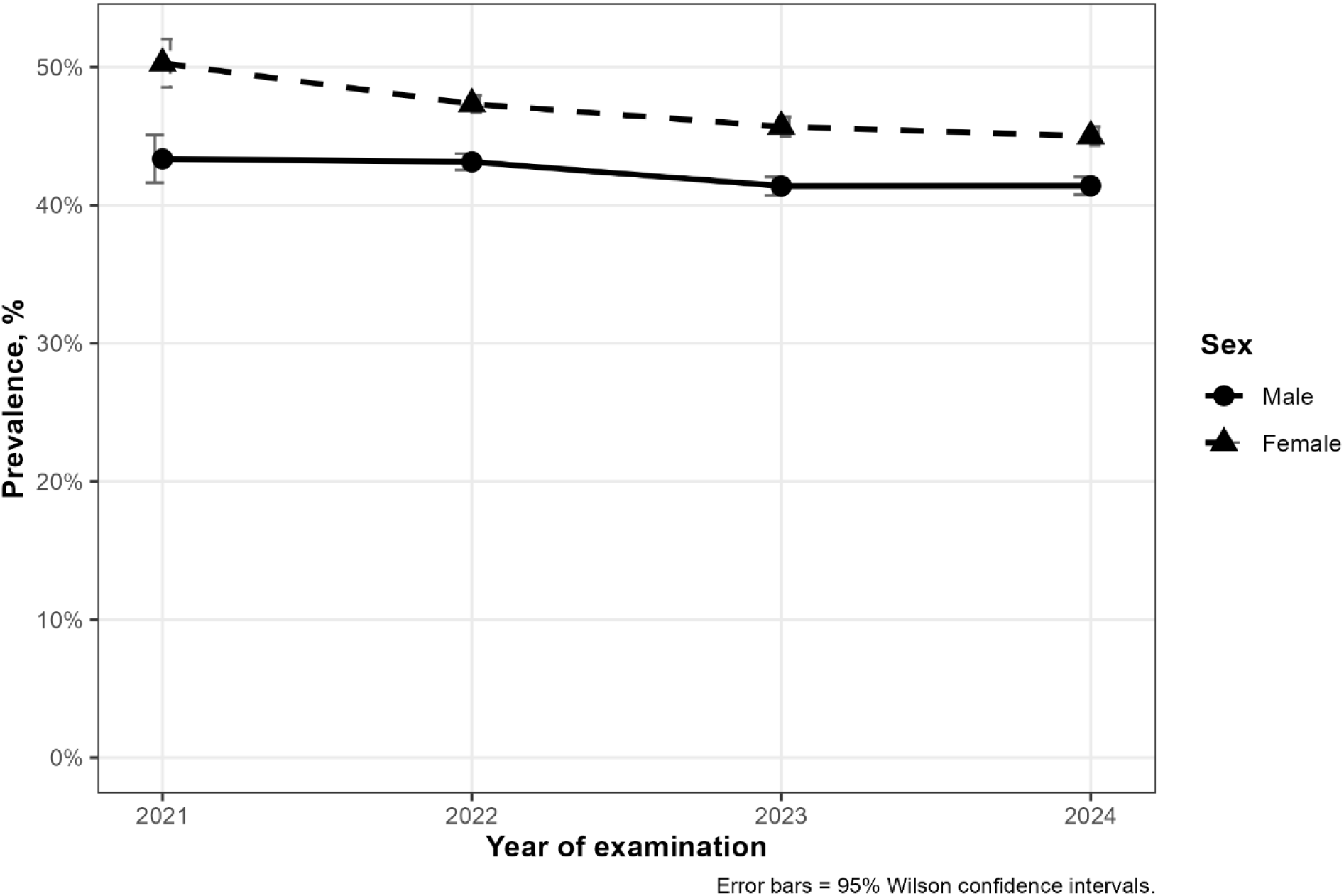
Sex-stratified prevalence of any refractive error by calendar year. Annual prevalence of any refractive error stratified by sex. Points indicate annual prevalence estimates and shaded bands represent 95% Wilson confidence intervals. Estimates were based on school vision screening data. RE = refractive error.

### Quantitative refractive error measurements in the Hanoi autorefractor sub-cohort

Objective autorefractor measurements were available for 19,788 examinations performed in Hanoi between 2022 and 2024. The prevalence of myopia increased markedly from 3.7% in 2022 to 11.0% in 2024 (*P* for trend < 0.001), whereas the prevalence of moderate/high myopia declined from 0.8% to 0.4% over the same period (*P* = 0.002). Conversely, the prevalence of hyperopia decreased substantially from 82.1% in 2022 to 61.0% in 2024 (*P* < 0.001). Astigmatism remained common throughout the study, affecting approximately two-thirds of children (67.0% to 71.4%), while anisometropia increased from 13.9% in 2022 to 20.4% in 2024 (all trend *P* < 0.001) (**Table 2**).

Consistent with these prevalence patterns, the mean spherical equivalent (right eye) declined from 0.80 ± 2.42 D in 2022 to 0.44 ± 1.34 D in 2024, indicating a progressive shift toward more myopic refractive status (**Table 4**).

**Table 4.**
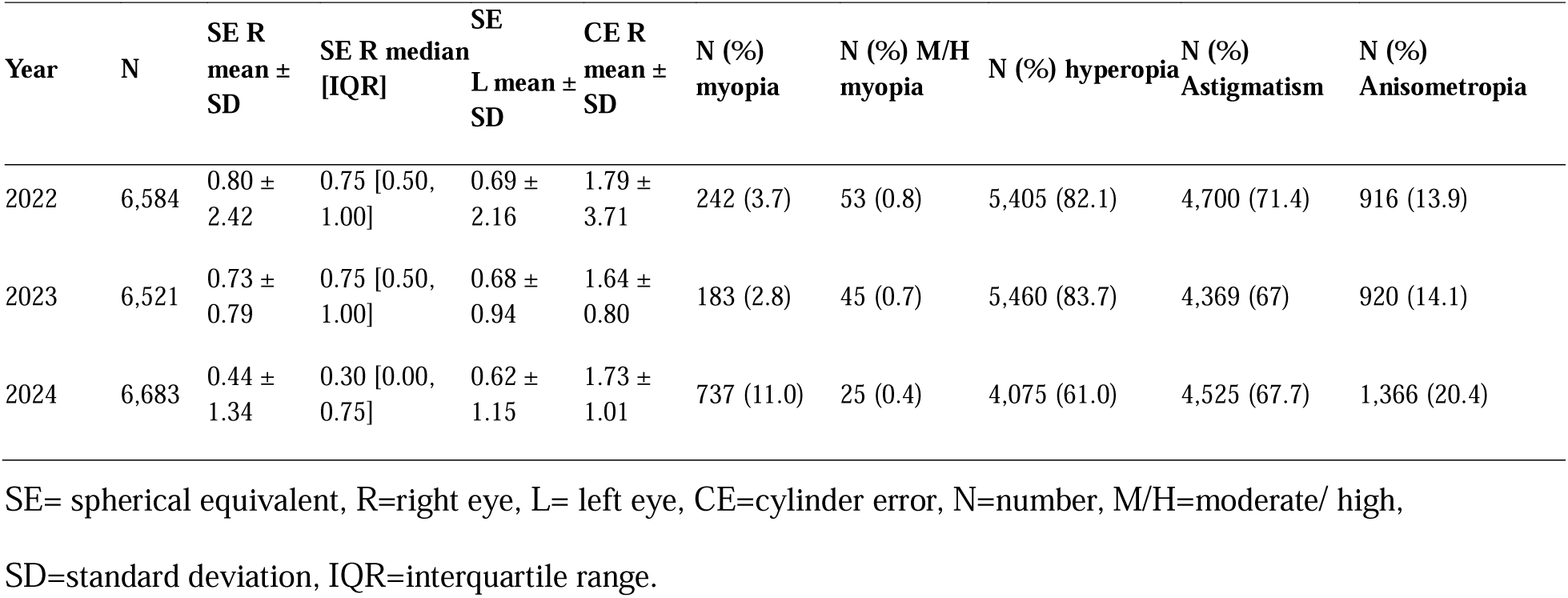
Quantitative refractive error measurements in the Hanoi autorefractor cohort by year, 2022 to 2024.

**Figure 7** further demonstrates marked differences in refractive error patterns across educational stages. Hyperopia predominated among kindergarten children throughout the study period, whereas myopia became increasingly common in older children. Astigmatism remained highly prevalent across all school levels with relatively modest temporal variation, while anisometropia showed a progressive increase in prevalence in successive school grades. These findings indicate an age-dependent transition from predominantly hyperopic to increasingly myopic refractive profiles during school years.

**Figure 7.**
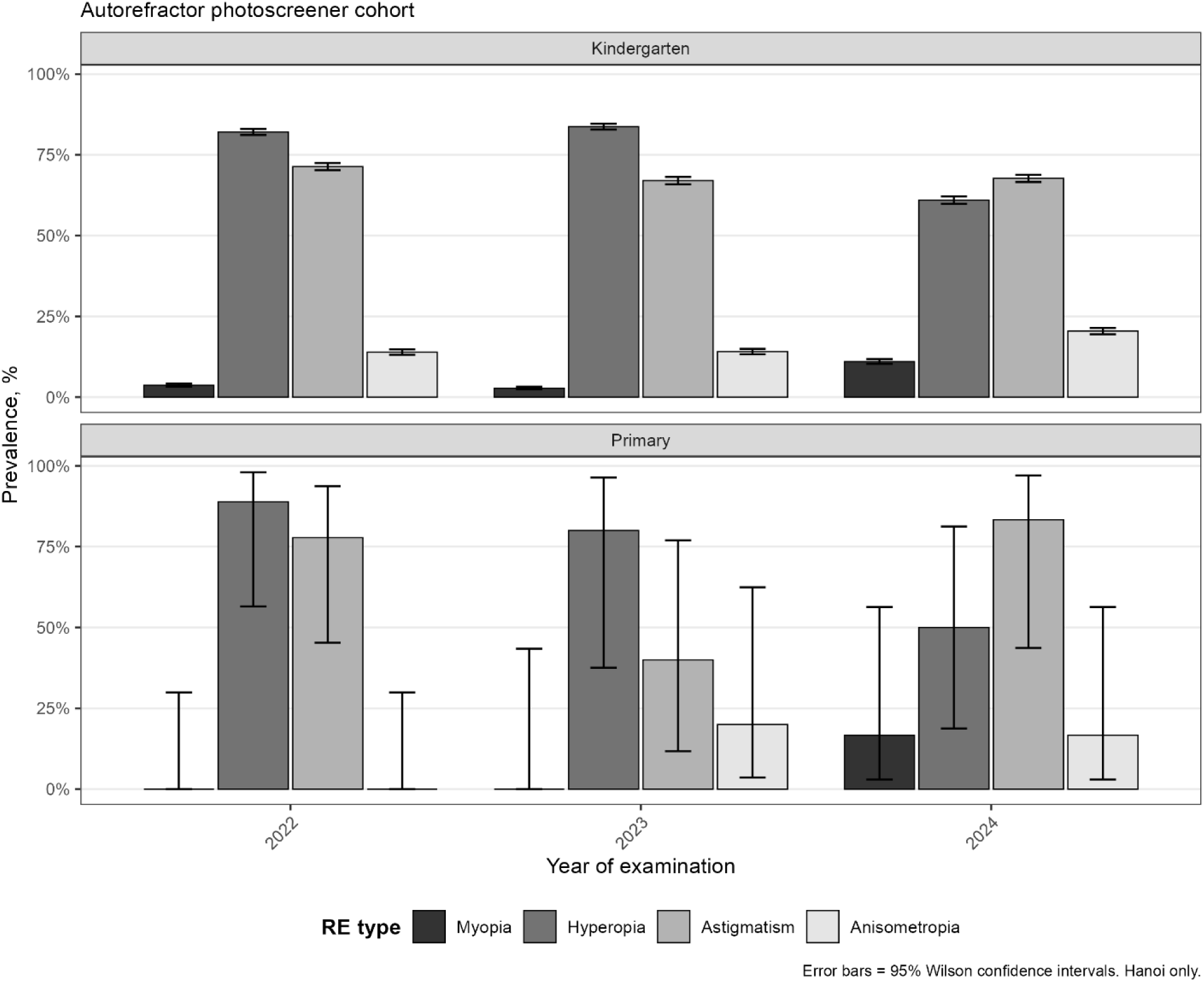
Refractory error type prevalence by school level and year for Hanoi autorefractor sub-cohort. Bar plots show the prevalence of myopia, hyperopia, astigmatism and anisometropia according to school level and calendar year among children examined using an autorefractor. Error bars indicate 95% Wilson confidence intervals. RE = refractive error.

## DISCUSSION

In this four-year, three-city school health screening program covering 141,339 examinations of children aged 3 to 18 years, over half of the children examined had some form of refractive error, with prevalence differing by city, sex, school level, and age. In the Hanoi autorefractor sub-cohort, where refractive error type could be classified objectively, hyperopia and astigmatism were far more common than myopia overall, though the proportion of children with confirmed myopia nearly tripled between 2022 and 2024, from 3.7% to 11.0%. This combination, a high overall burden alongside a rapidly rising myopia signal, sits at an interesting point within the wider Vietnamese and regional picture.

Our overall “any refractive error” prevalence estimates, drawn from routine school vision screening rather than dedicated eye examinations, were notably higher than the 37.6% pooled prevalence reported in a recent meta-analysis of Vietnamese schoolchildren^8^. This gap likely reflects differences in case definition and age range rather than a true difference in disease burden. School screening protocols typically flag any reduced or abnormal visual acuity for referral, which captures early hyperopia and astigmatism that would not meet stricter clinical thresholds, and our cohort also included children as young as three years, an age group in whom physiologic hyperopia is expected and who are largely absent from prior Vietnamese surveys, which have focused almost entirely on children six years and older^8^. Read this way, our findings do not contradict the existing literature so much as extend it downward in age, into a population where refractive development itself, rather than pathological myopia, still explains most of what is measured.

The rise in confirmed myopia within the Hanoi autorefractor cohort is, we think, the more clinically important signal in these data, and it fits comfortably within the broader East and Southeast Asian trajectory. Vietnamese secondary-school myopia prevalence has previously been shown to climb rapidly, from 13.1% in 2009 to 33.7% by 2014^8^, and similar accelerating trends have been documented across China^2,11^. What our data add is evidence that this rise may now be detectable at younger ages within a single urban Vietnamese cohort followed over just three years, a pattern consistent with observations from China that the mean age of myopia onset has been shifting earlier, from 10.6 years in 2005 to 7.6 years in 2021, alongside erosion of children’s physiologic hyperopic reserve^11^. Because earlier onset carries a substantially higher lifetime risk of high myopia and its associated complications^12^, the possibility that Vietnamese children are also beginning to show this shift deserves close monitoring, even though our study period is still short and autorefraction was only available in one of the three cities.

The marked differences we observed between cities also echo a consistent theme in the regional literature, that urban residence and its accompanying lifestyle, including reduced outdoor time and more intensive near work, is associated with higher refractive error prevalence^4,16^. Hai Phong and Hanoi showed higher adjusted prevalence than Ho Chi Minh City in our data, a pattern that may relate to differences in school screening practices, referral thresholds, or the age structure of children examined across sites rather than to a true difference in underlying eye health, and this is a limitation we cannot fully resolve with the present data. The higher prevalence we observed among girls is, however, consistent with several Vietnamese and Chinese studies^8,10^, and the rise in prevalence from primary through upper secondary school level mirrors the well-established pattern of myopia accumulating with age and years of schooling^4^.

This study has several strengths, including its large sample size, its coverage of three major cities, its multi-year design, and the availability of objective autorefraction data, which is uncommon in Vietnamese school health research. It also has important limitations. Refractive error in the full cohort was based on school vision screening rather than cycloplegic refraction, the gold standard for epidemiological studies, and non-cycloplegic assessment is known to overestimate myopia and underestimate hyperopia^2^. Missing data were also unevenly distributed, reaching nearly 18% in Hanoi in 2022, which could bias estimates if missingness was related to the outcome. Finally, because the autorefractor sub-cohort was restricted to Hanoi, we cannot confirm whether the myopia trend seen there is also occurring in the other two cities.

Taken together, these findings suggest that Vietnamese urban school health programs should look beyond overall refractive error prevalence and pay closer attention to trends in confirmed myopia at younger ages, since this is where the region’s rising burden has historically first become visible^2,11^. Expanding objective autorefraction screening beyond Hanoi, and building the kind of longitudinal, standardized surveillance that has so far been lacking in Vietnam^8^, would help clarify whether the myopia shift seen in this study reflects a genuine, citywide, or nationwide trend and would strengthen the evidence base needed to guide school-based vision screening and myopia prevention and control policy in Vietnam and similar countries.

## DECLARATIONS

### Ethical Statement

The study was approved by Vinmec Ethical Committee (approval number 0231/2024/CN/HDDD VMEC). Informed consent and written informed consent from the parent/guardian of participant under 18 years of age was waived for this study because this study performed retrospective analysis of de-identified data.

### Contributors

N.T.H. did conceptualization, data curation, formal analysis, investigation, methodology, project administration, resources, software, supervision, validation, visualization, writing original draft, and writing review & editing.

K.P.M.T. did data curation, formal analysis and writing review & editing.

All authors read and approved the manuscript.

### Declaration of Interests

All authors declare no competing interests.

### Data Sharing Statement

R code is available from the corresponding author upon reasonable request. Individual patient-level data cannot be shared due to applicable privacy regulations and the terms of the institutional ethics approval.

### Role of the funding source

This study did not receive any funding.

### Use of Artificial Intelligence

All scientific content, analyses, and interpretations are the original work of the authors. The authors used AI-assisted tools for language editing and grammar checking during manuscript preparation.

## Notes

### Competing Interest Statement

The authors have declared no competing interest.

