## Supplementary Materials for "Refractive error prevalence in urban Vietnamese children: a 2021 to 2024 school health analysis"

Table S1. Missing data in refractive error (RE) assessment by city and year

| **City** | **Year** | **n examined** | **n missing RE** | **% missing RE** | **N sex missing** | **N age missing** |
| --- | --- | --- | --- | --- | --- | --- |
| Hanoi | 2022 | 50,146 | 8,851 | 17.7 | 0 | 0 |
| Hanoi | 2023 | 28,970 | 1,144 | 3.9 | 0 | 0 |
| Hanoi | 2024 | 29,077 | 542 | 1.9 | 0 | 0 |
| Ho Chi Minh City | 2021 | 4,845 | 7 | 0.1 | 0 | 0 |
| Ho Chi Minh City | 2022 | 8,192 | 395 | 4.8 | 0 | 0 |
| Ho Chi Minh City | 2023 | 9,764 | 246 | 2.5 | 0 | 0 |
| Ho Chi Minh City | 2024 | 11,244 | 131 | 1.2 | 0 | 0 |
| Hai Phong | 2021 | 1,447 | 1 | 0.1 | 0 | 0 |
| Hai Phong | 2022 | 2,934 | 89 | 3.0 | 0 | 0 |
| Hai Phong | 2023 | 3,115 | 79 | 2.5 | 0 | 0 |
| Hai Phong | 2024 | 3,122 | 32 | 1.0 | 0 | 0 |

RE= refractive error.

Table S2. Stratified crude prevalence of refractive error (RE) outcomes.

| Outcome | Sex | n examined | n with outcome | Prevalence, % (95% CI) | Stratifier | School level | City |
| --- | --- | --- | --- | --- | --- | --- | --- |
| Any RE | Male | 58,262 | 30,911 | 53.1 (52.6, 53.5) | Sex |  |  |
| Any RE | Female | 53,779 | 31,451 | 58.5 (58.1, 58.9) | Sex |  |  |
| Impaired visual acuity | Male | 57,238 | 32,603 | 57.0 (56.6, 57.4) | Sex |  |  |
| Impaired visual acuity | Female | 52,263 | 27,594 | 52.8 (52.4, 53.2) | Sex |  |  |
| Myopia (HN) | Male | 10,629 | 639 | 6.0 (5.6, 6.5) | Sex |  |  |
| Myopia (HN) | Female | 9,159 | 523 | 5.7 (5.3, 6.2) | Sex |  |  |
| Hyperopia (HN) | Male | 10,629 | 8,014 | 75.4 (74.6, 76.2) | Sex |  |  |
| Hyperopia (HN) | Female | 9,159 | 6,926 | 75.6 (74.7, 76.5) | Sex |  |  |
| Astigmatism (HN) | Male | 10,629 | 7,422 | 69.8 (68.9, 70.7) | Sex |  |  |
| Astigmatism (HN) | Female | 9,159 | 6,172 | 67.4 (66.4, 68.3) | Sex |  |  |
| Any RE |  | 25,925 | 17,908 | 69.1 (68.5, 69.6) | School level | Kindergarten |  |
| Any RE |  | 41,665 | 13,701 | 32.9 (32.4, 33.3) | School level | Primary |  |
| Any RE |  | 30,410 | 19,823 | 65.2 (64.6, 65.7) | School level | Lower secondary |  |
| Any RE |  | 14,041 | 10,930 | 77.8 (77.1, 78.5) | School level | Upper secondary |  |
| Impaired visual acuity |  | 5,312 | 5,255 | 98.9 (98.6, 99.2) | School level | Kindergarten |  |
| Impaired visual acuity |  | 57,167 | 35,042 | 61.3 (60.9, 61.7) | School level | Primary |  |
| Impaired visual acuity |  | 33,678 | 14,988 | 44.5 (44.0, 45.0) | School level | Lower secondary |  |
| Impaired visual acuity |  | 13,344 | 4,912 | 36.8 (36.0, 37.6) | School level | Upper secondary |  |
| Myopia (HN) |  | 19,768 | 1,161 | 5.9 (5.6, 6.2) | School level | Kindergarten |  |
| Myopia (HN) |  | 20 | 1 | 5.0 (0.9, 23.6) | School level | Primary |  |
| Hyperopia (HN) |  | 19,768 | 14,925 | 75.5 (74.9, 76.1) | School level | Kindergarten |  |
| Hyperopia (HN) |  | 20 | 15 | 75.0 (53.1, 88.8) | School level | Primary |  |
| Astigmatism (HN) |  | 19,768 | 13,580 | 68.7 (68.0, 69.3) | School level | Kindergarten |  |
| Astigmatism (HN) |  | 20 | 14 | 70.0 (48.1, 85.5) | School level | Primary |  |
| Any RE |  | 76,539 | 47,138 | 61.6 (61.2, 61.9) | City |  | Hanoi |
| Any RE |  | 27,906 | 10,027 | 35.9 (35.4, 36.5) | City |  | Ho Chi Minh City |
| Any RE |  | 7,596 | 5,197 | 68.4 (67.4, 69.5) | City |  | Hai Phong |
| Impaired visual acuity |  | 76,787 | 37,178 | 48.4 (48.1, 48.8) | City |  | Hanoi |
| Impaired visual acuity |  | 23,934 | 20,474 | 85.5 (85.1, 86.0) | City |  | Ho Chi Minh City |
| Impaired visual acuity |  | 8,780 | 2,545 | 29.0 (28.0, 29.9) | City |  | Hai Phong |

RE=refractive error, HN=Hanoi autorefractor sub-cohort, 95%CI=95% confidence interval.
